# Reliability and competitiveness of a novel balance field test: A cross-sectional pilot study

**DOI:** 10.1101/2024.05.11.24307223

**Authors:** Gerald Jarnig, Reinhold Kerbl, Mireille N.M. van Poppel

## Abstract

Balancing tests in the field provide important information about the physical fitness and injury risk of athletes. These tests provide useful data on an athlete’s ability to maintain balance in dynamic and unforeseen circumstances, allowing for a more detailed assessment of their athletic performance. We have designed a novel balance test and tested main quality criteria and competitiveness for it. A total of 117 children (girls = 52.1%) aged 10.4 ± 0.5 years participated in the tests. The raw ABC scores show good to excellent test-retest reliability for all participants. Girls show a higher reliability (excellent) than boys (moderate to good). The comparison of raw data of the performance in the ABC and existing balance field tests shows strong correlation of the results. The test duration of the ABC is significantly (p<.001) shorter than that of existing balance field tests. The statistical and graphical analysis of the calculation of the normal distribution shows that the results of the ABC are closer to the normal distribution than those of the existing balance field tests.

The combination of high reliability and competitiveness makes the Austrian Balance Check an extremely attractive option for various areas of application. Overall, the novel designed test is a promising instrument that has the potential to significantly improve the standards of balance testing procedures in the field. The authors recommend a large-scale study to review the test quality criteria of the ABC, including gold standard comparison and development of age- and gender-specific reference values, in order to increase interest in implementing the Austrian Balance Check in international test batteries.

## Introduction

When testing fitness in children and adolescents, we differ between tests that are carried out in laboratories with expensive equipment and time-consuming procedures and field tests that are primarily organized in schools [1–4]. Some European countries show how a continuous nationwide fitness and health monitoring system can work [5–7], while in other countries, such as Austria, this is still not achieved. In Austria, for example, this is limited to projects and regional initiatives [8–12], but there is still no consensus on a nationwide, continuous and long-term system. The practical experience of the authors of this report has shown that many of these test systems used in Europe are, in particular, very time-consuming and cost-intensive and are therefore limited in their suitability for nationwide use (Table 1).

**Table 1.**
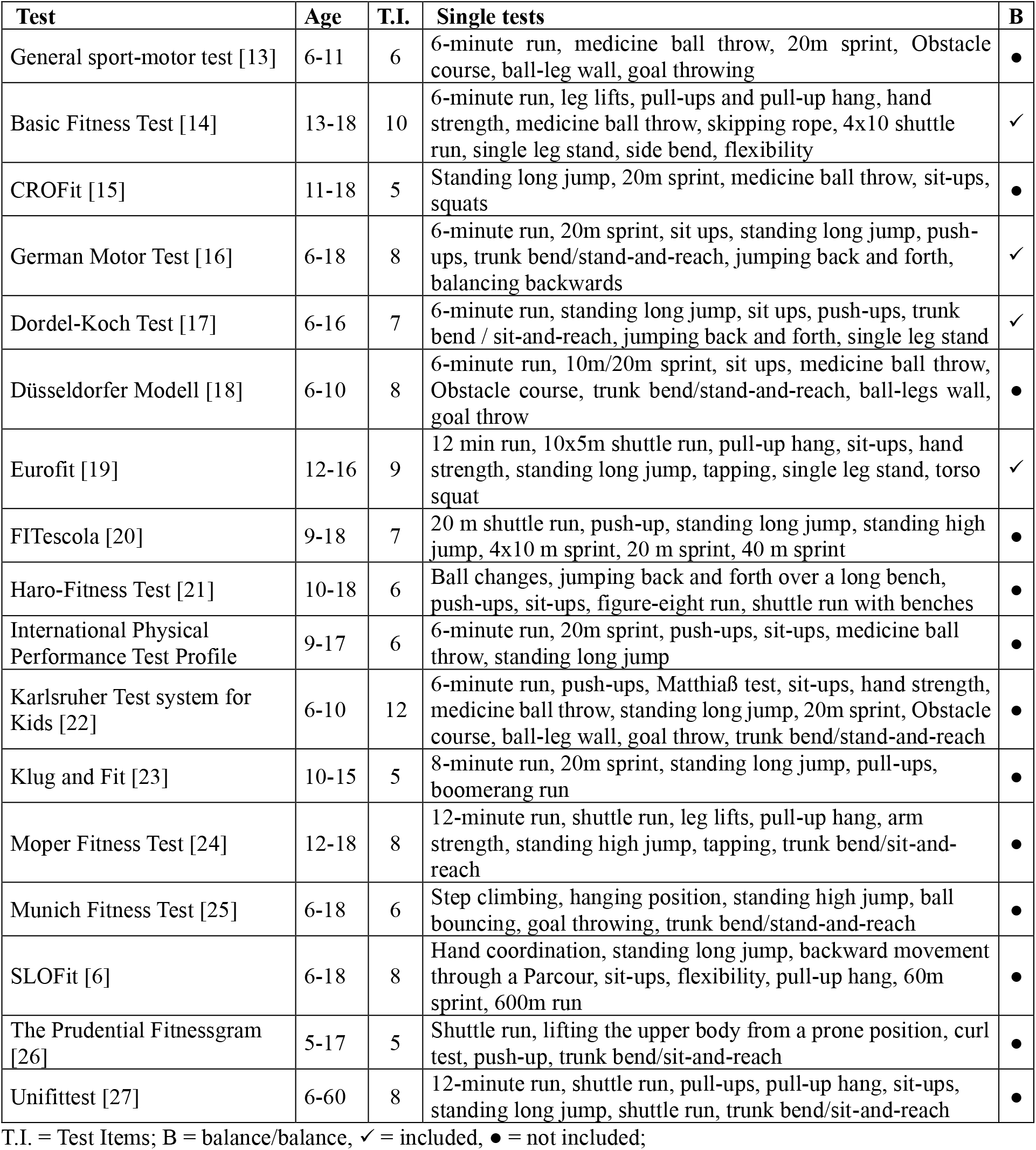
Established fitness monitoring systems in Europe.

For this reason, a project was started in 2019 to develop an Austrian Fitness and Health Test Battery (AUT FIT) [28] with three main components that would make it stand out from other test batteries.

The aim was to enable the testing of larger groups based on short time requirements, cost-effective implementation and simple evaluation of the results [28]. In the development process, many existing test systems were analyzed. Based on the information obtained and the practical experience of AUT FIT development, the authors became aware of an additional problem: balance testing. Existing balance field tests are either very time-consuming or involve high costs for the testing material (Table 2). Furthermore, it is noticeable that only a few established international test batteries include balance testing (see Table 1).

**Table 2.** Established fitness monitoring systems in Europe.

| Test | Especially expensive test equipment | Additional necessary materials |
| --- | --- | --- |
| General sport-motor test [13] |  | Target throwing system |
| Basic Fitness Test [14] | Hand dynamometer about 200 € / device | Dynamometer |
| CROFit [15] |  |  |
| German Motor Test [16] | Backward balancing system about 450 € / device |  |
| Dordel-Koch Test [17] | Classic Sit and Reach Box about 250 € / device |  |
| Düsseldorfer Modell [18] | Light barrier system about 600 € / device | Target throwing system |
| Eurofit [19] | Hand dynamometer about 200 € / device | Dynamometer, Tapping device |
| FITescola [20] |  |  |
| Haro-Fitness Test [21] |  |  |
| International Physical Performance Test Profile |  |  |
| Karlsruher Test system for Kids [22] | Hand dynamometer about 200 € / device | Dynamometer, Material for obstacle course, target throwing system |
| Klug and Fit [23] |  | Material for obstacle course |
| Moper Fitness Test [24] | Classic Sit and Reach Box about 250 € / device;<br>Hand dynamometer about 200 € / device | Dynamometer, Tapping device |
| Munich Fitness Test [25] |  | Target throwing system; |
| SLOFit [6] |  | Dynamometer, Material for obstacle course, target throwing system; |
| The Prudential Fitnessgram [26] | Classic Sit and Reach Box about 250 € / device |  |
| Unifittest [27] | Classic Sit and Reach Box about 250 € / device |  |

The aim of this work was to design a balance test aimed at testing large groups in a short time and at low cost, and to carry out first checks on reliability and competitiveness.

## Methods

Based on the basics of the test manual designed by the authors, a cross-sectional study was conducted in the greater Klagenfurt area to test the scientific suitability of the designed test manual. This was approved by the Research Ethics Committee of the University of Graz, Styria, Austria (GZ. 39/23/63 ex 2018/1/9).

### Selection of schools and participants

With the help of a random generator, 4 out of 12 primary schools from the greater Klagenfurt area were selected which participated at the same time in a multi-year health study (2019 to 2022 [10,11,29,30]). All of the school directors agreed to participate in the study. The following inclusion criteria were defined: The children had to be attending 4th grade in the 2021/22 school year and be able to perform all balance-specific tests without restrictions. In spring 2022, we invited all 147 children from the selected classes to participate. In total, 117 children took part in the reliability tests (Test-Re-Test) and 104 children took part in the competitiveness test (Table 3).

**Table 3.**
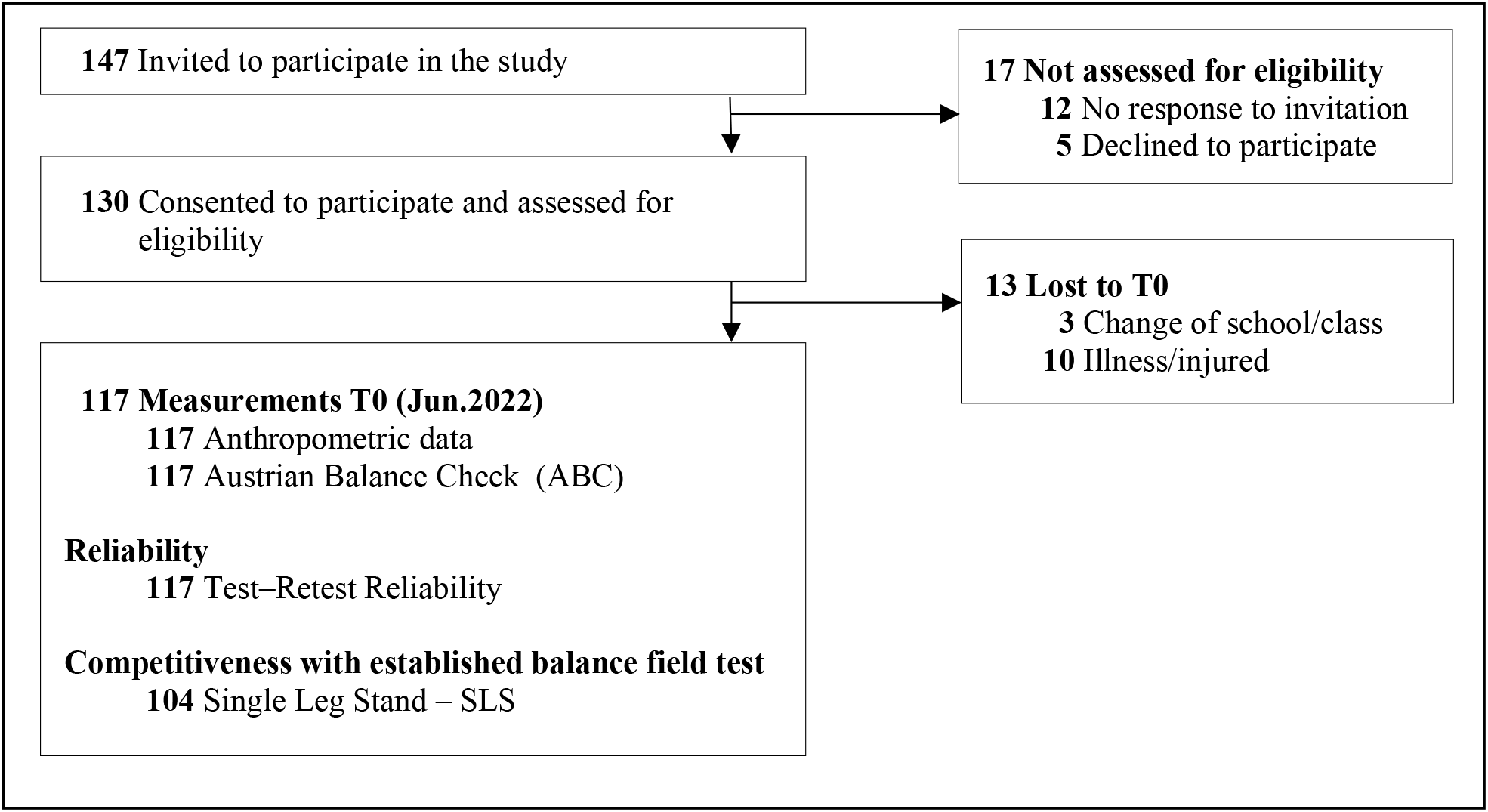
Flow diagram.

### Procedure

The measurement of anthropometric data and balance was carried out by trained members of the research team and took place in the schools during physical education (PE) lessons. All tests were conducted without shoes on a non-slip surface, with participants wearing standard sportswear.

### Anthropometry

Height (cm) was measured to the nearest 0.1 cm using a SECA 213 stadiometer and weight (kg) was measured to the nearest 0.1 kg using a Bosch PPW4202/01 body scale. The BMI was calculated by dividing the body weight by the height in meters squared.

### Austria Balance Check (ABC)

The Austrian Balance Check (ABC) is a three-stage field test, with increasing levels of difficulty, to assess static balance.

The participants stand with their preferred leg (supporting leg), the other leg ( playing leg) is moved forwards and upwards until the heel of the playing leg is at a height of about 5 cm above a floor marker, with both arms supported at the hips.

The test consists of three levels of difficulty; in order to be allowed to complete the next level, a previous level must be successfully completed. Each participant has a maximum of two attempts to successfully complete a level. A level is considered successfully completed when the maximum time available for a run has been reached. An attempt ends when the maximum time to complete a level has been reached, one hand loses hip contact, the standing leg leaves the mark on the floor, the playing leg is moved backwards, the eyes are opened in levels 2 and 3 or the chin moves forwards and downwards in level 3.

Participants collect 1 point per level for every 10 seconds of successful performance. If they successfully complete levels 1 and 2, a total of 7 points are given for each completed level. If a level is successfully completed on the first attempt, an extra point is given for each level. If a participant does not manage to successfully complete level 1 or 2, the points collected in both attempts of this level are added to the overall assessment. This also applies to level 3, where participants must complete both attempts, regardless of whether the maximum time to be completed was completed on the first attempt or not. A maximum of 20 points can be achieved in total. (detailed information on the test structure, the procedure, the documentation and the test evaluation of the ABC can be found in the supplementary material - Test Manual Austrian Balance Check (ABC))

### Existing balance tests carried out to check the Reliability and competitiveness

#### One-legged stand according to Jarnig (SLS JA21)

The children were instructed to stand with one leg on a thin wooden board with their hands on their hips and to hold this position for as long as possible, for a maximum of 45 seconds. The test was performed twice with the preferred leg and the best result (in seconds) for each leg was taken into account. If the maximum value (45 seconds) was reached on the first attempt, no second attempt was made. Exact details on the setup of the test device are given in Jarnig et al. 2021 [28]. The longest of the maximum of two trials is included in the overall evaluation. In addition, the time (in minutes) required to collect data for a group and the number of group participants are documented (Time SLS JA21).

#### One-legged stand according to Dordel [17](SLS DKT)

The children are instructed to stand on a skipping rope with their preferred leg (standing leg) and to keep the other leg (free leg) bent at the knee, upwards and in the air. Over a period of 60 seconds, there should be no or as little ground contact as possible (FC SLS DKT) with the free leg. If the child touches the ground with the free leg, the starting position should be restored as quickly as possible. The test was carried out with eyes open. The number of ground contacts is included in the overall score. In addition, the time (in seconds) required for a participant to collect data is documented (Time SLS - DKT).

### Weight classification

National reference values [31] were used for BMI standardization and weight classification. The national reference data were expressed in BMI centile curves (i.e. equicurves, called EQUI BMI in this report) [31]. The absolute BMI values were converted to EQUI BMI values according to the procedure described in Mayer et al. [31] (based on Cole et al. [32]). The EQUI-BMI curves were used to project actual BMI to cut-off values at age 18 years to categorize children’s weight into five categories (underweight < 18.5 kg/m2, normal weight = 18.5 to 25.0 kg/m2, overweight ≥ 25.0 kg/m2, obese ≥ 30.0 kg/m2, morbid obesity ≥ 35.0 kg/m2).

### Test quality criterion - reliability

In order to assess the test-retest reliability of the ABC, the children were tested twice by the same test administrator (interrater reliability), with an interval of between one and two weeks between the test days.

### Checking the competitiveness

In order to assess the competitiveness of the ABC, testing times of established balance field tests were compared with the duration of the ABC. In addition, the raw data of established balance field tests and the ABC were checked for normal distribution and the correlations of the raw scores of the individual tests were calculated.

### Statistical analysis

Continuous variables are expressed as mean (M) and standard deviation (SD), categorical variables as absolute value (n) and percentage (%) for descriptive statistics.

For test-retest reliability, a two-sided mixed intraclass correlation coefficient (ICC) was calculated on the basis of individual measures and absolute agreement for the raw scores of the ABC tests. To define reliability, the 95% CIs of the ICCs were interpreted as follows: 95% CI values below 0.5 were considered to indicate poor reliability, values between 0.5 and 0.75 were considered to indicate moderate reliability, values between 0.75 and 0.9 were considered to indicate good reliability and values above 0.90 were considered to indicate excellent reliability.

To evaluate differences in competitiveness, the test durations of established balance field tests [17,28] were compared using a paired t-test with the duration of the ABC. In addition, the data were statistically and visually checked for normal distributions.

The Spearman correlation coefficient (ρ) between the performance on the ABC and the results of other balance tests was calculated. The strength of the correlations was classified according to Cohen [33], with a weak correlation classified as ρ ≥0.1, a moderately strong correlation as ρ ≥0.3 and a strong correlation as ρ ≥0.5.

No imputation of the data was performed. All statistical analyses were performed in SPSS 29.0 (IBM SPSS Statistics 29, IBM, New York, USA) with a significance level of p < 0.05.

## Results

A total of 117 children (girls = 52.1%) aged 10.4 ± 0.5 years participated in the tests (Tables 3 & 4). In the weight classifications, the percentage of girls (91.8%) who were underweight or normal weight was higher than that of boys (67.9%) (Table 4).

**Table 4.**
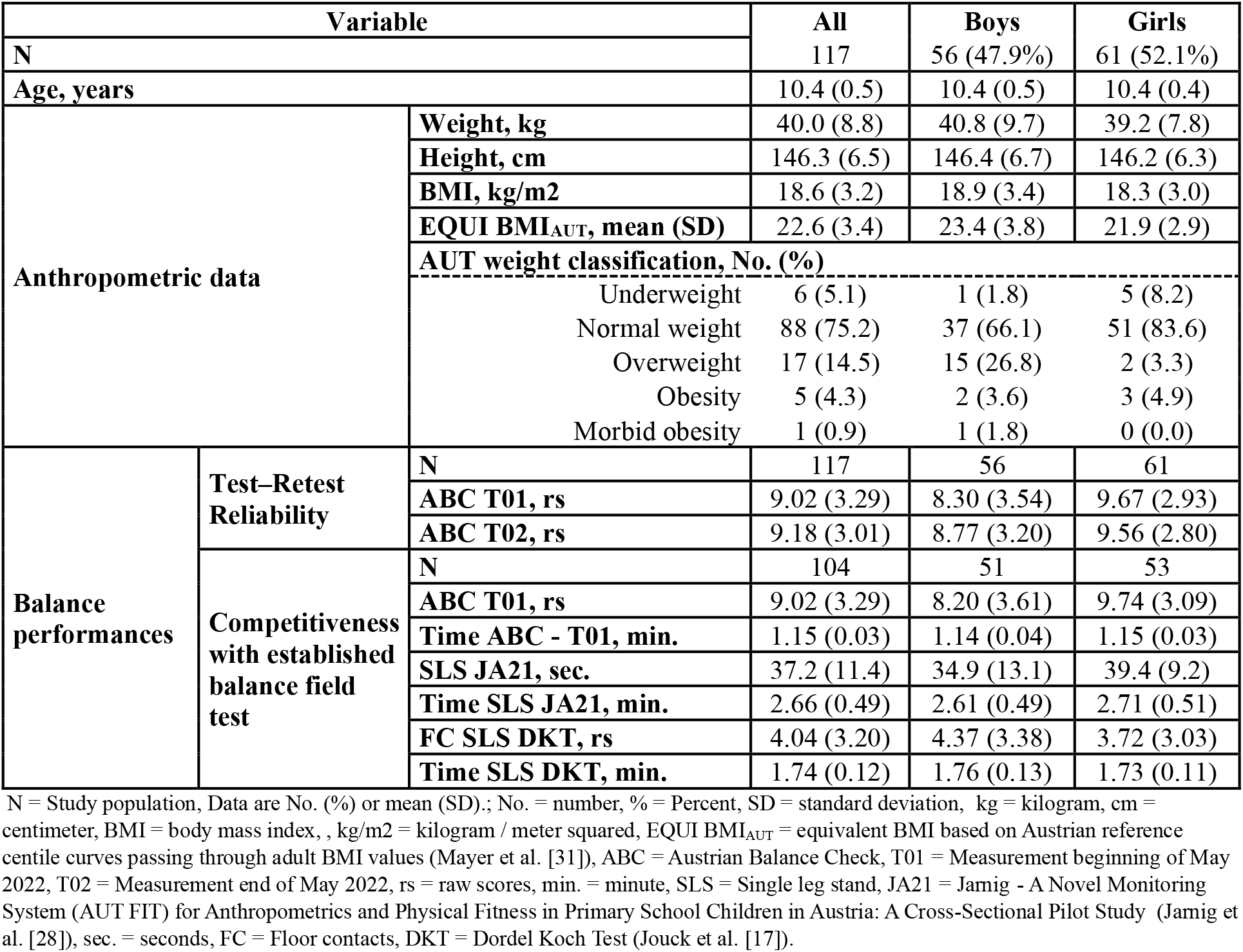
Overall sample characteristics.

The raw ABC scores show good to excellent test-retest reliability for all participants. Girls show a higher reliability (excellent) than boys (moderate to good) (Table 5).

**Table 5.** Intraclass correlations between test and retest scores from the Austrian Balance Check.

| Variable | ICC | 95% CI |  | Agreement |
| --- | --- | --- | --- | --- |
|  |  | Lower | Upper |  |
| All | 0.894 | 0.848 | 0.927 | Good to Excellent |
| Boys | 0.804 | 0.667 | 0.885 | Moderate to Good |
| Girls | 0.987 | 0.979 | 0.992 | Excellent |

Explanation: To determine reliability (test [ABC T01] – retest [ABC T02]), the 95% CIs of the ICCs were interpreted as follows: 95% CI values below 0.5 were considered to indicate poor reliability, values between 0.5 and 0.75 were considered to indicate moderate reliability, values between 0.75 and 0.9 were considered to indicate good reliability, and values above 0.90 were considered to indicate excellent reliability. ICC = intraclass correlation, CI = confidence interval, ABC = Austrian Balance Check, T01 = Measurement beginning of May 2022, T02 = Measurement end of May 2022.

The test duration of the ABC is significantly (p<.001) shorter than that of existing balance field tests (Table 6).

**Table 6.** Comparison of testing times for different balance field tests by using paired t-tests.

| Pairwise comparisons | Paired Differences |  | t | df | P value | E.s. |
| --- | --- | --- | --- | --- | --- | --- |
|  | Mean diff. (95% CI) | S.E. |  |  |  |  |
| **Time ABC T01 vs. Time SLS JA21** | -0.60 (-0.62 to -0.57) | 0.01 | -48.035 | 103 | *** | L |
| **Time ABC T01 vs. Time SLS DKT** | -1.52 (-1.62 to -1.42) | 0.05 | -31.135 | 103 | *** | L |

Data shows the mean difference in testing times measured in minutes per participant; Mean diff = mean difference, CI = confidence interval, SE = standard error, t = test statistic, df = degrees of freedom, P value means = \*\*\* $p < .001$ ; E.s. = Effect size, L = large effect according to Cohen ( $d \geq 0.8$ ). ABC = Austrian Balance Check, T01 = Measurement beginning of May 2022, SLS = Single leg stand, JA21 = Jarnig - A Novel Monitoring System (AUT FIT) for Anthropometrics and Physical Fitness in Primary School Children in Austria: A Cross-Sectional Pilot Study (Jarnig et al. [28]), FC = Floor contacts, DKT = Dordel Koch Test (Jouck et al. [17]).

The statistical and graphical analysis of the calculation of the normal distribution shows that the results of the ABC are closer to the normal distribution than those of the existing balance field tests (Figure 1 & Table 7).

**Table 7.** Checking for normal distribution of data from the balance field tests by using the Shapiro-Wilk test.

| Variable |  | Shapiro-Wilk |  |  |
| --- | --- | --- | --- | --- |
|  |  | t | df | P value |
| **ABC T01** | **All** | 0.971 | 104 | 0.023 * |
|  | **Boys** | 0.967 | 51 | 0.17 |
|  | **Girls** | 0.968 | 53 | 0.16 |
| **SLS JA21** | **All** | 0.719 | 104 | <.001 *** |
|  | **Boys** | 0.767 | 51 | <.001 *** |
|  | **Girls** | 0.662 | 53 | <.001 *** |
| **FC SLS DKT** | **All** | 0.89 | 104 | <.001 *** |
|  | **Boys** | 0.902 | 51 | <.001 *** |
|  | **Girls** | 0.879 | 53 | <.001 *** |

Data shows Results of the Shapiro-Wilk test for checking the normal distribution; t = test statistic, df = degrees of freedom, P value means = \* $p < .05$ , \*\* $p < .01$ , \*\*\* $p < .001$ ; ABC = Austrian Balance Check, T01 = Measurement beginning of May 2022, SLS = Single leg stand, JA21 = Jarnig - A Novel Monitoring System (AUT FIT) for Anthropometrics and Physical Fitness in Primary School Children in Austria: A Cross-Sectional Pilot Study (Jarnig et al. [28]), FC = Floor contacts, DKT = Dordel Koch Test (Jouck et al. [17]).

**Figure 1.**
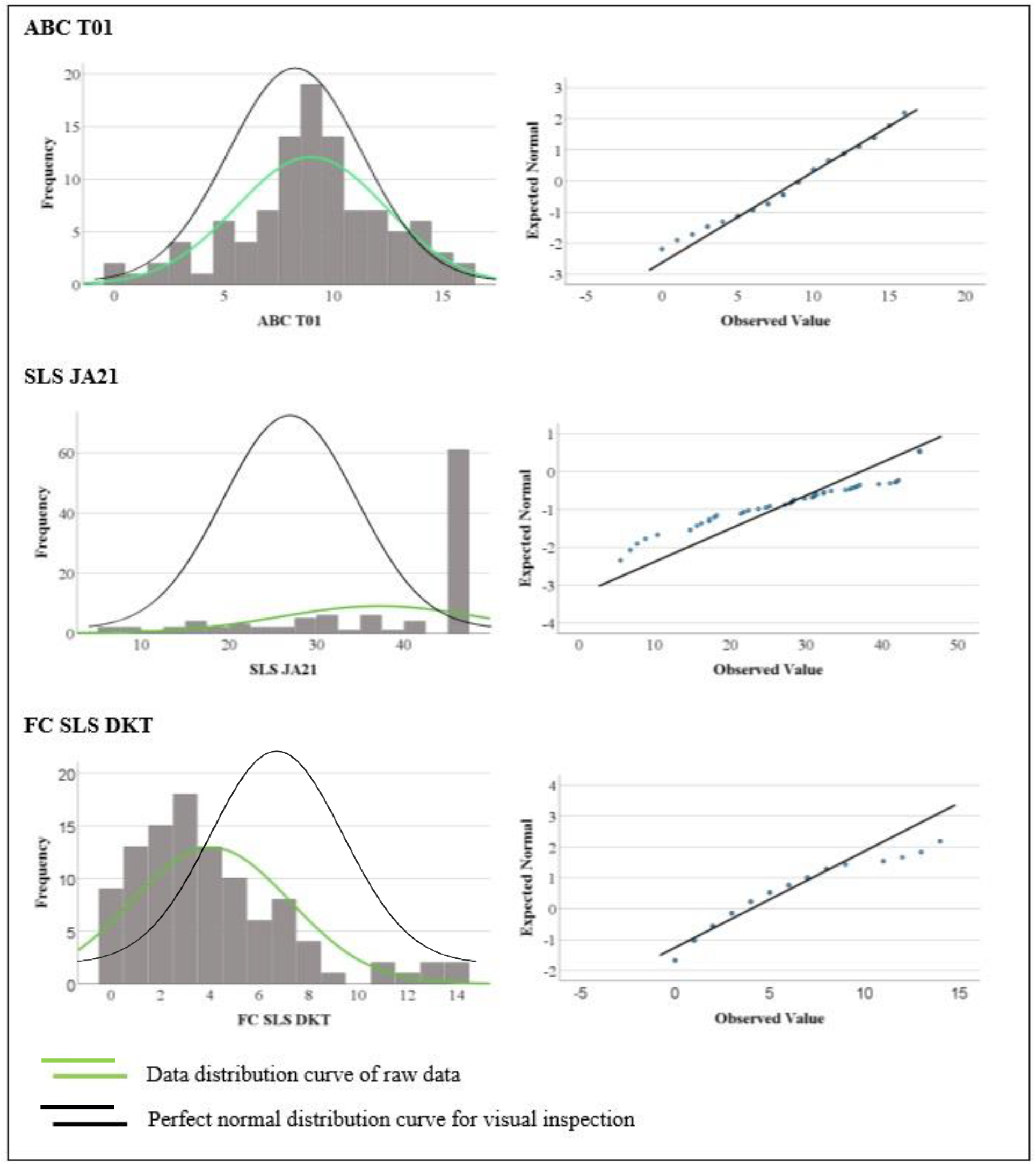
Graphical checking of the normal distribution using histograms and Q-Q plots for the total study population comparing competitiveness with established balance field tests ABC = Austrian Balance Check, T01 = Measurement beginning of May 2022, SLS = Single leg stand, JA21 = Jarnig - A Novel Monitoring System (AUT FIT) for Anthropometrics and Physical Fitness in Primary School Children in Austria: A Cross-Sectional Pilot Study (Jarnig et al. [28]), FC = Floor contacts, DKT = Dordel Koch Test (Jouck et al. [17]).

The comparison of raw data of the performance in the ABC and existing balance field tests shows strong correlation (Table 8) of the results.

**Table 8.**
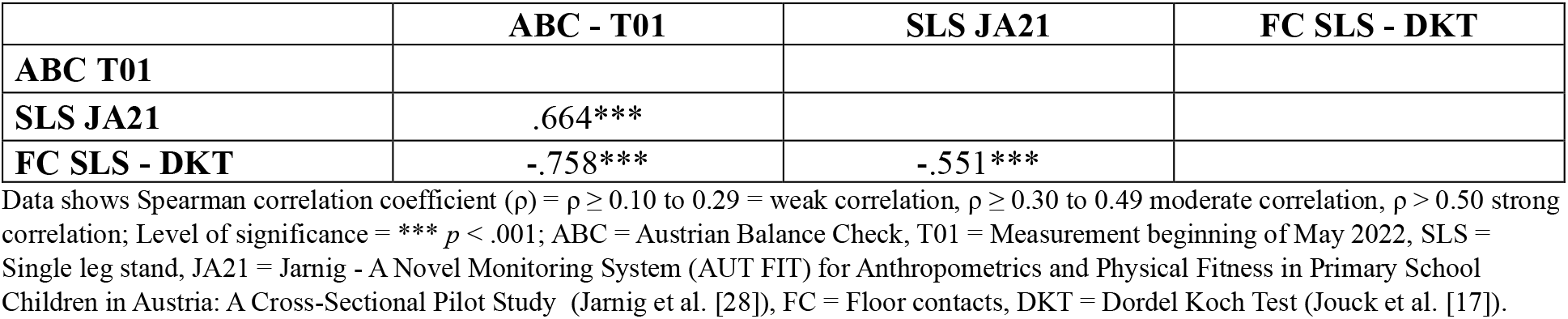
Correlation among results of different static balance tests assessed by Spearman’s correlation coefficient.

|  | **ABC - T01** | **SLS JA21** | **FC SLS - DKT** |
| --- | --- | --- | --- |
| **ABC T01** |  |  |  |
| **SLS JA21** | .664*** |  |  |
| **FC SLS - DKT** | -.758*** | -.551*** |  |

Data shows Spearman correlation coefficient ( $\rho$ ) = $\rho \geq 0.10$ to $0.29$ = weak correlation, $\rho \geq 0.30$ to $0.49$ moderate correlation, $\rho > 0.50$ strong correlation; Level of significance = \*\*\* $p < .001$ ; ABC = Austrian Balance Check, T01 = Measurement beginning of May 2022, SLS = Single leg stand, JA21 = Jarnig - A Novel Monitoring System (AUT FIT) for Anthropometrics and Physical Fitness in Primary School Children in Austria: A Cross-Sectional Pilot Study (Jarnig et al. [28]), FC = Floor contacts, DKT = Dordel Koch Test (Jouck et al. [17]).

## Discussion

The newly developed balance field test is characterized by a remarkably high reliability, in this context the newly designed test has shown an extremely reliable measurement of the variables to be tested, which clearly underlines its quality and reliability. In addition, the competitiveness of the test compared to other established methods is confirmed. A notable advantage of the ABC is its efficiency in terms of test duration. Compared to existing tests, it takes significantly less time (p<.001) to complete without sacrificing quality or reliability.

A shorter test duration offers a number of advantages that can have a positive impact on various aspects of the testing process and its application. For both test participants and test administrators, a shorter test duration means direct time savings. This is particularly important in situations where time is a limited resource, such as in schools, personnel employment or clinical practice. Reduced test duration can therefore indirectly lead to lower overall costs, as less time and resources are required to carry out the test. This can result in significant savings, especially for large-scale or repeated testing. Long test durations can lead to tester tiredness, which can have a negative impact on the quality of performance and the reliability of test results. Test participants are more willing to participate in a test if they know that it has a short duration.

Overall, shorter test duration and the associated benefits can help to improve the efficiency, accuracy and acceptability of test batteries, increasing their effectiveness in different application areas.

### Strengths and limitations

The major strength is the design of an innovative field test that demonstrates high reliability and competitiveness. However, there are still limitations due to the small study population with a minimal age range and the lack of verification of objectivity and validity.

## Data Availability

All data produced in the present work are contained in the manuscript

## Availability of data and materials

The data presented in this study are available upon request from the corresponding author. The data are not publicly available due to privacy/ethical restriction.

## Abbreviations

AUT FIT: Austrian Fitness and Health Test Battery
SDS: standard deviation scores
BMI: body mass index
ABC: Austrian Balance Check
SLS: Single leg stand

## Acknowledgments

This study was organized by the non-profit association NAMOA—Nachwuchsmodell Austria. The authors would like to thank all participants and their guardians; the trainers and staff of this study; Wolfgang Modritz for the initiation of this study; None of the individuals listed were financially compensated.

## Funding

This research was funded by the Austrian Federal Ministry for Arts, Culture, Civil Service, and Sport, grant number GZ2021-0.361.671.

## Author information

### Author Contributions

Conceptualization, G.J. and M.N.M.v.P.; methodology, G.J.; formal analysis, G.J.; investigation, G.J.; resources, G.J.; data curation, G.J.; writing—original draft preparation, G.J.; writing—review and editing, G.J., R.K., and M.N.M.v.P.; visualization, G.J.; supervision, M.N.M.v.P.; project administration, G.J.; funding acquisition, G.J. All authors read and agreed to the published version of the manuscript.

## Ethics declarations

### Ethical approval and consent to participate

This study was conducted according to the guidelines of the Declaration of Helsinki and approved by the Research Ethics Committee of the University of Graz, Styria, Austria (GZ. 39/23/63 ex 2018/19). All participants gave their consent to participate in the study.

### Consent for publication

Not applicable.

### Competing interests

The authors declare no conflicts of interest. The funders had no role in the design of the study; in the collection, analyses, or interpretation of data; in the writing of the manuscript, or in the decision to publish the results.

### Additional information

Additionally, information’s are given in the supplementary materials.

## Notes

### Competing Interest Statement

The authors have declared no competing interest.

### Author Declarations

This was approved by the Research Ethics Committee of the University of Graz, Styria, Austria (GZ. 39/23/63 ex 2018/1/9).

